# COVID-19-related disruption and resiliency in immunisation activities in LMICs: a rapid review

**DOI:** 10.1101/2023.06.12.23291133

**Authors:** Anna-Maria Hartner, Xiang Li, Katy Gaythorpe

## Abstract

**Objectives:** Rapid review to determine the extent that immunisation services in LMICs were disrupted by the COVID-19 pandemic and what factors can be considered to build resilience in future.

**Setting:** We searched PubMed on 28th Feb 2023 for studies published after 1st December 2019 in English that focused on LMICs.

**Participants:** Screening and data extraction were conducted by two experienced reviewers with one reviewer vote minimum per study per stage. Of 3801 identified studies, 66 met the eligibility criteria.

**Outcomes:** Routine vaccine coverage achieved; Supplementary immunisation activity timing; Vaccine doses given; Timing of vaccination; Supply chain changes; factors contributing to disruption or resilience.

**Results:** Included studies showed evidence of notable declines in immunisation activities across LMICs related to the COVID-19 pandemic. These have included reductions in achieved routine coverage, cancellation or postponement of campaigns, and underimmunised cohorts. Immunisation was most disrupted in the early months of the pandemic, particularly March to May 2020; however, the amount of recovery seen varied by country, age-group, and vaccine. Though many countries observed partial recovery beginning after lockdown policies were lifted in 2020, disruption in many countries has also continued into 2021. It has also been noted that clinician staff shortages and vaccine stock outs caused by supply chain disruptions contributed to immunisation delays but that concern over COVID transmission was a leading factor. Key resiliency factors included community outreach and healthcare worker support. Finally, whilst our search took place in February 2023, the latest dataset used across all studies was from November 2022 and many focused on 2020; as a result some of the study conclusions do not take recovery into account.

**Conclusions:** There is limited information on whether reductions in vaccination coverage or delays have persisted beyond 2021. Further research is needed to assess ongoing disruptions and identify missed vaccine cohorts.

**Strengths and limitations of this study:**

- The rapid synthesis of findings related to immunization disruption and recovery to-date allows for key insights to target missed cohorts and identify research gaps.
- We include a narrative analysis of disruption across LMICs; this review benefits from the inclusion of barriers, enablers, and resilience to/in service provision.
- The search strategy was limited to studies published on PubMed up to February 28th, 2023, meaning not all relevant research meeting inclusion criteria may have been captured.

## 1 Introduction

The COVID-19 pandemic began on December 12th 2019 and quickly spread globally, adding to the strain on existing healthcare provision and creating unique problems in terms of service delivery [1]. Throughout 2020 there were disruptions to screening for cancer, maternal health services, care for chronic conditions such as diabetes, and immunisations [2]. This strain on health services has continued past 2020, as even those that have to recovered pre-COVID levels of visits and surveillance have to catch-up missed cohorts and delayed treatments.

LMICs disproportionately bear the burden of vaccine preventable diseases [3]; however, globally vaccination has seen a stagnation in coverage and zero-dose children are a concern. The issue of zero dose or underimmunised children is particularly important as it can hint at wider heterogeneity in healthcare access which may have been exacerbated by the pandemic [4]. It is estimated that 67 million children missed vaccinations between 2019 and 2021; of those, 48 million were zero-dose children [5]. Furthermore, targeting zero-dose children can be more difficult as they are often in harder-to-reach areas, particularly in LMICs where 1 in 6 children living in rural areas are zero-dose [5].

Resilient healthcare systems can withstand additional and unusual strains whilst maintaining priority services. Yet, is is still uncertain what factors contributed to disruption or resilience in light of the COVID-19 pandemic, which was a unique test on global healthcare systems. These factors and considerations may be instrumental in preparing for future healthcare strains such as those potentially caused by other epidemics, climate change, or antimicrobial resistance. As such, understanding the key factors for disruption due to the COVID-19 pandemic is critical for future planning in order to minimise the negative consequences of disruptions.

In order to understand the current state of vaccination coverage disruption, and highlight factors contributing to resilience, we undertook a rapid review of the existing literature. This focused on LMICs as they bear the majority of burden of vaccine preventable diseases. We included studies that not only discuss the quantitative measures of disruption such as reduced immunisation coverage and cancelled campaigns, but also more qualitative discussions of the factors contributing to disruption or characteristics of resilient systems.

## 2 Aim and research questions

The aim of this review was to understand the extent of disruptions in vaccination coverage due to the COVID-19 pandemic and what factors contributed to the disruption or resilience. Specifically, our research questions were:

RQ1: To what extent were immunisation services in LMICs disrupted by the COVID-19 pandemic?

RQ2: How did disruption vary by geography, demography or socioeconomic group?

RQ3: What factors contributed to coverage disruption or resilience?

## 3 Methods

A rapid review (RR) was conducted using streamlined systematic review methods and reported in accordance with the Preferred Reporting Items for Systematic Reviews and Meta-Analyses (PRISMA) guidelines [6].

### 3.1 Procedure

We searched PubMed on 28th Feb 2023 for studies published after 1st December 2019 in English with search terms (((COVID-19) OR (SARS-CoV-2))) AND (immunisation OR vaccination) AND (disruption OR delay* OR postpon*). Studies were included if they focused on disruption to vaccination activities due to the COVID-19 pandemic in LMICs. Studies were excluded if they focused on high income countries only, examined disruption due to other factors ie. not related to the pandemic, or were reviews, commentaries or modelling studies without novel data.

### 3.2 Study selection, data extraction and quality assessment

Search results were imported into the Covidence (www.covidence.org) systematic review management tool where duplicates were removed. Titles and abstracts were screened by one reviewer, full text review was completed by two reviewers with conflicts resolved through consensus.

Each study was extracted by one reviewer into a Google sheet. We extracted information on i) last date of included data, ii) countries studied, iii) qualitative findings related to the research questions RQ1, RQ2 and RQ3, and iv) binary data on whether routine immunisation, SIAs, doses, schedule timing or supply chains were mentioned in the study. A second reviewer was consulted where there was uncertainty concerning the extracted data.

To expedite the review we did not use a formal quality assessment tool. instead, we focused on the scope of the study in terms of population, schedules considered and time window to assess the generalisability of the findings.

### 3.3 Synthesis

There were two main types of evidence to synthesise. Quantitative information on percentage drops in coverage achieved, doses administered or SIAs postponed, and more qualitative discussion on contributing factors informed by surveys or questionnaires. We grouped results by research question with the first question the most quantitative. Finally we collate characteristics of the studies themselves such as countries studied or dates of included data, for which we have prepared summary statistics.

### 3.4 Patient and public involvement

There was no patient or public involvement in this study.

## 4 Results

### 4.1 Characteristics of studies

We found 3801 studies where 66 met the inclusion criteria 1. The majority of studies were published in either 2021 (n = 30; 45.45%) or 2022 (n = 27; 40.91%), though most studies only reported on data from 2020 (n = 46; 69.70%). 10 studies (15.15%) included data during the first 6 months of 2021; a further 8 (12.12%) included data between July and December of 2021. Only 2 studies (3.03%) included data from 2022; the most recent of these covered data through November of 2022.

Most (n = 16; 24.24%) of the studies considered multiple LMICs. Of those that only considered one country, India (n = 8; 12.12%), Ethiopia (n = 5; 7.58%), Brazil (n = 4; 6.06%) and Pakistan (n = 4; 6.06%) were the most frequently studied. The African continent was the most represented.

Most (n = 47; 71.21%) studies examined the effect of the COVID-19 pandemic on routine immunisation coverage, with an additional 6 (9.09%) reporting pandemic effects on supplementary immunisation activities. The change in the number of administered doses (n = 13; 19.70%) or the timing of doses (n = 11; 16.67%) was also reported by several studies; 7 (10.61%) reported disruptions in the vaccine supply chain.

**Figure 1:**
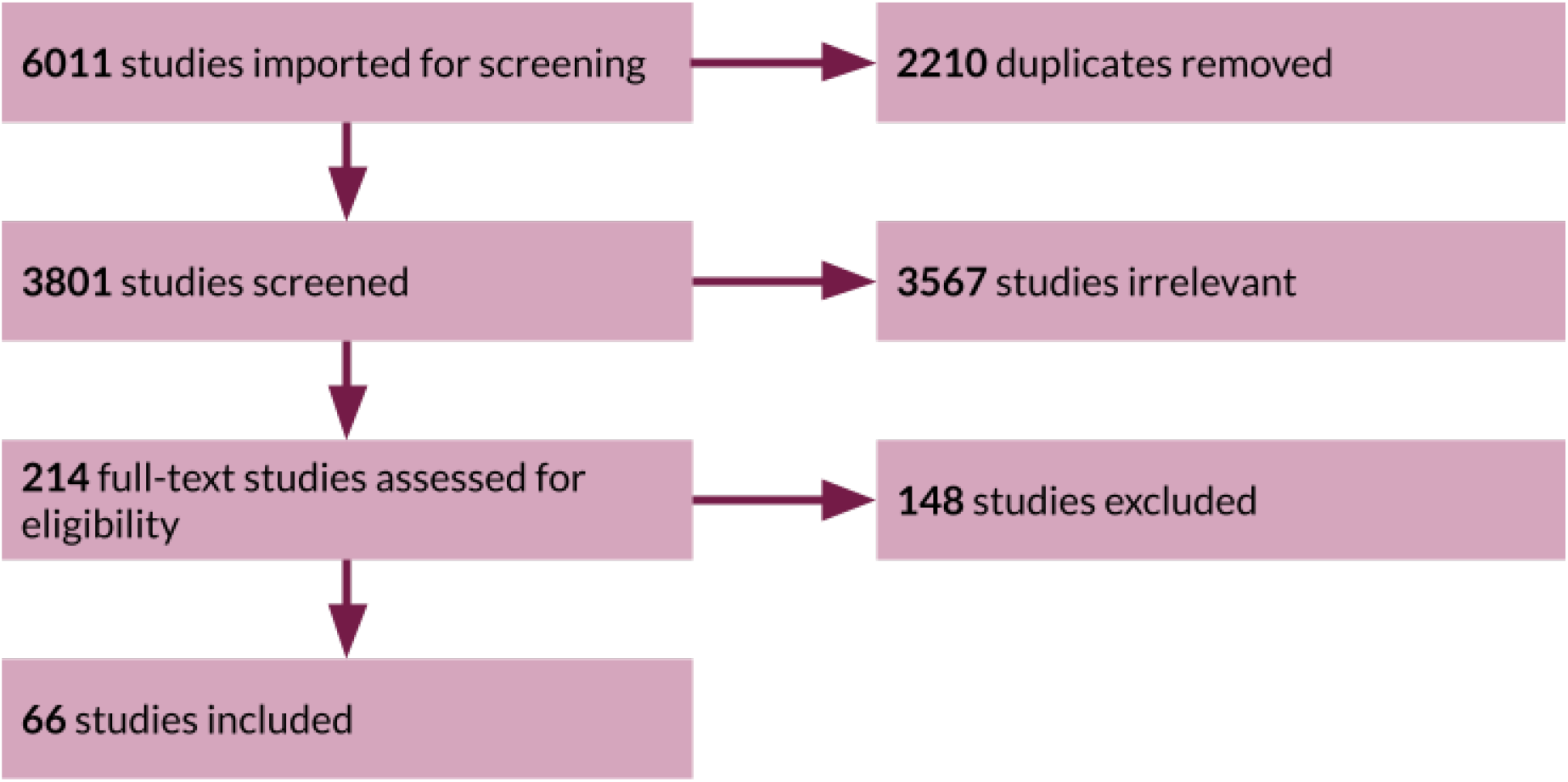
PRISMA flow of study selection. PRISMA, Preferred Reporting Items for Systematic Reviews and Meta-Analyses.

### 4.2 Extent of disruption

We divide this into a few main areas. Firstly, supply chains and vaccine availability, then the delivery of routine immunisation both in doses given and delays, then we examine supplementary immunisation activities, and finally, signs of recovery.

#### 4.2.1 Vaccine supply

Following the declaration of COVID-19 as a pandemic, there was a reduction of vaccine sales and periods of stockout and low availability of vaccines in some countries [7, 8, 9]. Vaccine sales between April and August 2020 fell by 9.5% [10] but some losses were recouped by catch-up activities [8].

#### 4.2.2 Routine immunisation

We divide insight by WHO region or country. In the WHO African Region there was a varied picture of disruption. In Ethiopia there were minimal disruptions up to August 2020 [11, 12, 13, 14]. Similarly, in DRC, disruptions in Kinshasa were minimal up to December 2020 [15] and in Kenya immunisation services were largely unaffected [16, 17, 18]. However, in South Africa full immunisation dropped in the first months of the pandemic, especially in April where it dropped by 30% [19, 20]. Ghana [21, 22, 23], Nigeria [24], Uganda [8], Liberia [25] and Sierra Leone [26] all saw drops in coverage in 2020 and whilst some countries had begun to see recovery in coverage achieved, this was not enough to compensate for missed cohorts [27]. In the WHO region of the Americas, there were declines in coverage reported for Dominican Republic, Mexico, Ecuador and Brazil; Dominican Republic saw a drop of 10 percentage points [28], vaccinations reduced by 36% in Mexico [29, 30], there were 14% fewer doses administered in Ecuador [31], and in Brazil approximately 20% of children missed vaccinations with a 18% overall decline in dose administered in the first year of the pandemic [32, 33, 34]. Although one study found no significant evidence of COVID-19 isolation measures on vaccines per child in Brazil [35].

In the Eastern Mediterranean WHO region, drops in coverage were seen for Lebanon, Afghanistan, Jordan and Pakistan [36, 37, 38, 39, 40] of 31%, 21%, 6-16% and 30-48% respectively over the initial stages of the pandemic.

In the South East Asian WHO region, there were significant disruptions [41]. In India, there were substantial drops in coverage across the majority of districts (88% [42]) especially in lockdown and early in the pandemic [43, 44, 45, 46]; as a result children born in India after COVID-19 had a 2-10% lower probability of timely vaccination compared to earlier cohorts [47]. In Nepal and Bangladesh, the most severe disruptions were also seen earlier in the pandemic, particularly in Bangladesh where 20-25% of planned outreach immunisation was cancelled between April and May 2020 [48, 49].

In the WHO European region, in Armenia, there were only small declines in coverage achieved [50]. Globally, there were substantial drops in routine immunisations in 2020 [9]. Overall, it was an estimated that there were 31% fewer vaccine doses given [51], in middle-income countries 14% of individuals delayed or missed vaccinations in the first 6 months of the pandemic [52], and there was a 20% increase on children who had not completed the 3-dose DTP series [53]. Whilst disruption varied by vaccine [54, 55], most saw the most severe declines in the 6 months of the pandemic followed by variable recovery [56] which may affect control and elimination efforts [57].

It was not only the total number of doses administered that was affected, but also when those doses were given. In China and India, the majority of interviewed caregivers delayed vaccination [58, 59, 60, 47, 52] and in Ecuador and Sierra Leone this delay was worse for last doses [31, 26].

#### 4.2.3 Supplementary immunisation activities

Overall, we found fewer studies focusing on supplementary immunisation activities (SIAs) or campaigns specifically; however, there are comprehensive records kept by the WHO campaign tracker as part of the immunisation repository [61]. SIAs were more disrupted in the early stages of the pandemic with 57% of planned campaigns globally postponed or cancelled because of COVID-19 by May 2020 [61]. By December 2020, this had fallen to 26% and many campaigns were reinstated from July 2020 onwards; by December 2021 this had fallen again to 16% of scheduled campaigns delayed or cancelled [61]. Overall, of those campaigns disrupted between March 2020 and December 2021, 59% had been reinstated [61]. Factors leading to postponement or cancellation of SIAs included non-pharmaceutical interventions [62] and stockouts or increased demand for general healthcare supplies [63, 8]. Additionally, whilst some SIAs had been reinstated, and there were plans for catchup activities, there are still large missed cohorts [44, 64, 57, 9].

#### 4.2.4 Recovery

Information on recovery is limited by the date ranges of the included studies which mainly focused on 2020 and 2021. A key finding is that while there were signs of improvement in routine immunisation coverage achieved and reinstated vaccination campaigns, there was not the positive increase needed to catch up missed cohorts [27, 51, 55, 49, 30, 65, 26]. It was also noted that pre-COVID levels of coverage had not been reached in many countries by the end of 2022 [57].

### 4.3 Heterogeneity in disruption

Heterogeneity in immunisation disruption was found across several factors, including geography, demography, wealth, and education; these are further detailed below. Variations in the extent of disruption by antigen were similarly reported in several studies [31, 37, 41, 10, 60].

#### 4.3.1 Geographic Heterogeneity

Despite significant decreases in immunisation in LMICs, there was significant geographic heterogeneity in the extent of disruption and in the regions and/or individuals affected. On a national level, several studies reported differences in the extent of disruption as a result of economic income classification [52, 41, 53] by WHO Region [63, 54, 53], by global burden of disease super-region [51], or by Gavi eligibility [53], with greater pandemic impact observed in low- and middle-income countries compared to high-income countries, affecting the primarily African Region, the Americas, and Asia. The reverse trend was seen for vaccine sales early in the pandemic (i.e. April to August 2020), with high-income countries experiencing a 20% decline and low-income countries observing a 10% increase [10].

On a sub-national level, many countries observed statistically significant differences between regions and provinces in regards to the change in health service utilisation [66, 30, 42] or routine immunisation coverage [31, 37, 46]. In some countries, certain provinces reported increases in immunisation service provision or doses for some vaccines, such as in the Southern Province of Rwanda, where measles and rubella immunisation increased [66]. Geographic heterogeneity was also observed in the subsequent recovery of services [42, 20].

While some countries reported differences in disruption between urban and rural areas, there was significant heterogeneity in the extent of disruption. One study found that the odds of immunisation in Ethiopia were higher in rural areas [11], while another observed greater initial declines in urban and periurban areas in South Africa, followed by recovery in these areas and declines in rural areas as the pandemic progressed [20]. In Pakistan, lockdown affected rural areas more than urban areas[39]. Geographic heterogeneity was also observed between Ethiopia’s hospitals and health centers, in which vaccine-related supplies were twice as likely to be affected by COVID-19 in hospitals [7], while in India children residing in “COVID-19 red zones” were more likely to face immunisation disruption [46]. Similarly, a study on polio outreach services in 33 African and Eastern Mediterranean countries found services necessary for “reaching their most vulnerable populations” were partially or severely disrupted [63].

#### 4.3.2 Demographic Heterogeneity

Few studies focused on the effects of demographic heterogeneity on COVID-19 related immunisation disruption, including factors such as gender, age, birth order, or caste. Only two studies looked at differences by gender; one found greater declines in females than males, though this decline was not significant [37]. The second, conducted in Brazil, also found no significant differences, but did find that infants were less likely to experience immunisation disruptions or delays compared to one-year old children[32]. This finding was similar two studies, conducted in Eastern India and in China, where increasing age of the child was found to be associated with immunisation delays [59, 60]. A study conducted in South-East Asia and the Western Pacific found similar results, in which early-infancy was less disrupted than infancy, school-entry age, and adolescent immunisation [41]. However, greater disruption was seen among infants compared to adult/elderly immunisation [41]; additionally, one study in Jordan found that children older than 12 months were less likely to experience delays [38].

Only two studies stratified results by maternal or caregiver age; one finding that increasing maternal age was associated with delayed vaccination [59], the other finding no association[38].

Additionally, one study conducted in China found firstborn children were less likely to experience delays [60], while another paper in India examined heterogeneity as a result of ethnicity or caste, finding lower castes had lower likelihoods of full immunisation and greater immunisation disruption, though these findings were not significant [46].

#### 4.3.3 Socioeconomic Heterogeneity

Contributors to socioeconomic heterogeneity in immunisation disruption largely included measures of household income and education. Two studies, one in Brazil and the other in India, found that missed vaccine doses were more likely in children from poorer households; in India it was additionally found that there were greater declines in immunisation among poorer subgroups [46, 32]. A study in South Africa found mixed results, finding declines in full immunisation and first dose of measles greater in wealthier quintiles at the start of the pandemic, but with faster positive recovery and continued declines among poorer subgroups as the pandemic progressed [20]. Only one study focused on education, similarly finding higher probability of incomplete immunisation and greater declines in households without formal education [46].

### 4.4 Factors contributing to coverage disruption and resilience

We divide this section into three key areas: health system barriers, vaccine demand and hesitancy, and resilience.

#### 4.4.1 Health System Barriers

Many of the initial challenges in maintaining immunisation services in LMICs were the result of health system and supply barriers during the early stages of the pandemic. Many countries reported issues with vaccine supply delays or stockouts [30, 7, 67, 65, 25, 8, 9, 68] and lack of personal protective equipment (PPE) for healthcare workers (HCWs), including masks, gloves, and other drugs and supplies [7, 67, 25, 24, 69, 42, 68, 44, 45, 42]. Disruption caused by vaccine stockouts or supplies was found to vary by WHO region [9] or by geographical sub-region [24, 69, 41]; notably one study in Southeast Asia and the Western Pacific found vaccine stockouts to be among the least important reasons for service provision delays [41]. A lack of logistical support impacting routine services or outreach, such as a lack of fuel or water, was reported by three studies in the WHO African region [67, 25, 24].

Similarly, HCW availability posed a significant challenge, with countries citing difficulties due to the diversion of staff to COVID-19 response, staff illness, and transportation difficulties, among others [25, 9, 24, 68, 44, 45, 24, 42]. One study in Kenya further reported disruption due to a HCW strike from December 2020 to January 2021 [18]. On an individual level, HCWs reported that pandemic-related stigma, stress, or fears impacted service delivery [7, 24, 48, 45, 68, 42], with some additionally reporting harassment by law enforcement or by patients themselves[24, 42]. Only one study, conducted at a tertiary health centre in Ghana, found no disruptions to vaccine supply or in HCW availability [22].

COVID-19 lockdowns and restrictions also resulted in cancelled immunisation services, clinic closures, or reduced healthcare access or services available[7, 70, 25, 24, 39, 41, 52, 46, 62, 38], with some reporting difficulties maintaining COVID-19 prevention rules, such as social distancing, due to non-compliant patients or a lack of space[67, 24, 45].

Competing priorities also meant some countries faced declines in funding for immunisation services or supplies, resulting in financial constraints [44, 69].

#### 4.4.2 Vaccine Demand and Acceptance

Many of the challenges in maintaining routine immunisation services during the COVID-19 pandemic also resulted from declining vaccine demand and increasing vaccine hesitancy among caregivers. Declines in vaccine demand were frequently attributed to travel barriers or difficulties in reaching immunisation services or clinics [46, 60, 14, 25, 9, 13, 45, 44, 48, 41], COVID-19 restrictions or requirements, including testing requirements, mask requirements, or lockdowns, [70, 67, 38, 40, 68, 48, 52], and financial constraints [13, 44, 48, 52]. One study, conducted in South East Asia and the Western Pacific, reported that while affordability issues contributed to immunisation service utilisation, it was among the lowest ranked reasons [41]. Some caregivers additionally reported low or no awareness of the availability of immunisation services, often believing clinics and hospitals were closed for routine immunisation services [67, 44, 22, 59].

Declines in vaccine demand due to fears of contracting COVID-19 at clinics or hospitals was pervasive, and one of the most reported causes across several studies [46, 52, 41, 48, 68, 44, 40, 13, 22, 45, 38, 9, 59, 67, 25]. Many others reported additional fear or stigma against healthcare providers, including fears that staff might be infected by the virus [22, 48, 45, 25, 68]. One survey of 100 caregivers at a tertiary health centre in Eastern India found that 83% of respondents agreed that “safety [was] more important than vaccination” [59]. Further unspecified declines in vaccine demand were noted by several studies [9, 65, 39].

Vaccine hesitancy factors were less commonly reported; misinformation and misbeliefs contributed to declines in demand in just two studies [9, 48], while fears specifically about vaccine side effects were found in just one study in a tertiary hospital in North Ghana [22]. One additional study in Liberia reported declines due to vaccine conspiracies, where parents believed their children would be injected with COVID-19 [25].

#### 4.4.3 Resiliency

Though few papers highlighted resiliency factors or enablers to immunisation during the COVID-19 pandemic, two key focuses included the community outreach to address declining vaccine demand and acceptance and the importance of improved healthcare worker support to increase service provision. In Jordan and China, alternative arrangements for childhood vaccination (i.e. outside of the standard service provision within healthcare clinics) was found to be key to maintaining immunisation demand, though in Jordan this insight was based on a survey of caregiver beliefs[38, 58]. Similarly, a community intervention highlighting the importance of maintaining timely vaccination, despite the pandemic, was crucial in Jordan and in Ethiopia [38, 13]. Ethiopia additionally reported decreased fear of COVID-19 as an enabling factor [13]. In India, adequate access to PPE, overcoming barriers to transportation for HCWs, community and/or family support, and training on COVID-19 management was crucial support HCWs in maintaining immunisation service provision[68]. Similarly, proactive communication and coordination on all levels of the healthcare system was essential in Ethiopia in maintaining health system resiliency[69].

## 5 Discussion

Despite the challenges faced by health systems during the COVID-19 pandemic, the WHO has continued to emphasise the importance of routine immunisation, noting that the last effects of immunisation declines can lead to higher burdens of disease and/or excess deaths[71]. This review highlights the extent of disruption faced by LMICs, finding significant heterogeneity between and within regions, countries, and individual demographics, but nevertheless showing declines in routine immunisation in 2020 and 2021 that had not often not recovered to pre-COVID levels.

SIAs and campaigns were postponed with few regions reporting full recovery. Many LMICs rely on outreach services to reach vulnerable populations, especially where access to health clinics or services are limited[9]. COVID-19 response efforts or mitigation strategies, including lockdowns, resulted in additional disruption to transportation services, logistical support, or supplies, often hindering additional outreach activities and limiting the services that were available. This has resulted in a deepening of existing coverage inequalities, with studies noting greater disruptions among households with lower incomes, formal education, or those situated in informal housing or in some regions, rural areas, emphasising the heterogeneity that existed prior to the pandemic [72].

The findings in this study are limited by the data available — the majority of studies utilised data from 2020, limiting much of our understanding of how routine immunisation services have recovered since countries lifted lockdown or other COVID-19 response policies. Our study also does not include grey literature, only articles, with the search limited to one database. Nevertheless, this study expands upon the findings of a systematic review of available literature on childhood disruptions to immunisation using data from 2020, which included 39 studies and found an overall median decline of 10.8% [73]. Our study highlights the findings through 2022 and emphasises the ongoing heterogeneity in immunisation, alongside the barriers and enablers to service provision.

Our findings emphasise the urgency required to target individuals and cohorts who may have missed out on routine immunisation or campaigns during the COVID-19 pandemic, ensuring the barriers highlighted by staff and caretakers, including low staff or service availability, vaccine or supply stockouts, and transportation barriers are mitigated. Importantly, approaches to combat fears, misinformation, or misbeliefs, including those surrounding COVID-19 transmission and risk, are critical. Though few studies touched on vaccine hesitancy, declining vaccine acceptance has become a formative issue, and additional strategies are required to prevent additional backsliding[74].

Rebuilding immunisation services in LMICs will require a greater focus on healthcare resilience, so that the disruption caused by future epidemics or disasters on routine immunisation services is minimal, and that recovery and performance rapid and improved through an adaptation to real-world events[75]. Many of the countries that showed service delivery resilience during the COVID-19 pandemic highlighted the need for proactive and ongoing communication and coordination across multiple interconnected systems, especially between the community and healthcare system. One study, published in May of 2023, offers an updated framework to address the idea of epidemic-ready primary healthcare. Importantly, this framework offers solutions to many of the observed barriers found in this review, focusing on adequate training, compensation, and protection for HCWs, reliable logistic and supply-chain infrastructure, and linkages to the community[76]. Given the reliance on primary health care and outreach systems for immunisation in LMICs, this approach may be a beneficial starting point, though notably, it will require a shift in how healthcare currently interacts with public health, alongside strong political commitment and financing[76]. Further research will be required to understand how post-pandemic disruption and recovery in immunisation services has progressed, especially in regards to vulnerable communities.

## Data Availability

All data produced in the present work are contained in the manuscript.

## 6 Acknowledgements

This work was carried out as part of the Vaccine Impact Modelling Consortium (www.vaccineimpact.org), which is jointly funded by the Bill & Melinda Gates Foundation (Grant Numbers INV-034281 and INV-009125 / OPP1157270) and Gavi, the Vaccine Alliance. The views expressed are those of the authors and not necessarily those of the Consortium or its funders. The funders were given the opportunity to review this paper prior to publication, but the final decision on the content of the publication was taken by the authors.

AMH, XL, and KAMG also acknowledge funding from the MRC Centre for Global Infectious Disease Analysis (reference MR/R015600/1), jointly funded by the UK Medical Research Council (MRC) and the UK Foreign, Commonwealth Development Office (FCDO), under the MRC/FCDO Concordat agreement and is also part of the EDCTP2 programme supported by the European Union; and acknowledge funding by Community Jameel.

